# T cell activation via the CD40 ligand and transferrin receptor and deficits in T regulatory cells are associated with major depressive disorder and severity of depression

**DOI:** 10.1101/2023.05.03.23289312

**Authors:** Muanpetch Rachayon, Ketsupar Jirakran, Pimpayao Sodsai, Atapol Sughondhabirom, Michael Maes

## Abstract

Major depressive disorder (MDD) is associated with T cell activation (Maes et al. 1990-1993), but no studies have examined the combined effects of T cell activation and deficits in T regulatory (Treg) cells on the severity of acute phase MDD. Using flow cytometry, we determined the percentage and median fluorescence intensity of CD69, CD71, CD40L, and HLADR-bearing CD3+, CD4+, and CD8+ cells, and cannabinoid type 1 receptor (CB1), CD152 and GARP-bearing CD25+FoxP3 T regulatory (Treg) cells in 30 MDD patients and 20 healthy controls in unstimulated and stimulated (anti-CD3/CD28) conditions. Based on cytokine levels, we assessed M1 macrophage, T helper (Th)-1, immune-inflammatory response system (IRS), T cell growth, and neurotoxicity immune profiles. We found that the immune profiles (including IRS and neurotoxicity) were significantly predicted by decreased numbers of CD152 or GARP-bearing CD25+FoxP3 cells or CD152 and GARP expression in combination with increases in activated T cells (especially CD8+CD40L+ percentage and expression). MDD patients showed significantly increased numbers of CD3+CD71+, CD3+CD40L+, CD4+CD71+, CD4+CD40L+, CD4+HLADR+, and CD8+HLADR+ T cells, increased CD3+CD71+, CD4+CD71+ and CD4+HLADR+ expression, and lowered CD25+FoxP3 expression and CD25+FoxP+CB1+ numbers as compared with controls. The Hamilton Depression Rating Scale score was strongly predicted (between 30-40% of its variance) by a lower number of CB1 or GARP-bearing Treg cells and one or more activated T cell subtypes (especially CD8+CD40L+). In conclusion, T helper and cytotoxic cell activation coupled with lowered Treg homeostatic defenses are key components of MDD and contribute towards greater immune responses and consequent neuroimmunotoxicity.

## Introduction

Depression is a major mental health concern that is becoming a more widespread global issue. In 2015, the World Health Organization estimated that approximately 4.4% of the world’s population, or approximately 322 million people, were affected by depression (1). Additionally, this report found that a similar percentage of the Thai population, or approximately 2.8 million people, suffers from depression (1).

There is now evidence that major depressive disorder (MDD) and bipolar disorder (BD) are associated with immune-inflammatory pathway activation. Studies have demonstrated that inflammatory markers, such as cytokines, chemokines, complement factors, and acute phase reactants, are elevated in depressed individuals (2-6). Both MDD and BD are characterized by activation of the immune-inflammatory response system (IRS) as indicated by elevated levels of pro-inflammatory cytokines, such as interleukin (IL)-1β, IL-6, IL-8 (CXCL8), IL-12, interferon (IFN)-γ, and tumor necrosis factor (TNF)-α (6), and the compensatory immune-regulatory system (CIRS), which prevents hyperinflammation by downregulating the IRS and producing anti-inflammatory cytokines, such as IL-4, IL-10 and transforming growth factor (TGF-β) (6). Under physiological conditions, IRS and CIRS interact in a balanced manner (6). During acute mood episodes, such as depressive episodes and mania, this balance is disrupted, resulting in an increase in the production of IRS and CIRS cytokines and a new IRS/CIRS setpoint (6-8). This is significant because certain IRS and CIRS cytokines have neurotoxic effects that may cause functional injury to neuronal brain circuits, particularly neuronal and astroglial projections (9).

As is the case with IRS/CIRS cytokines, there is evidence that imbalances in the number of T helper (CD4+) and T cytotoxic (CD8+) T cells and T regulatory (Treg) cells may contribute to the development of severe mood disorders (10-12). Flow cytometry-based immunophenotyping is a method for identifying activated T cells that contribute to IRS and CIRS profiles by producing pro- and anti-inflammatory cytokines. This technique enables us to distinguish between various categories of activated T effector (Teff) and Treg cells. Activated CD4+ and CD8+ cells that express activation markers such as CD69+, CD71+, CD40L+ (CD154+), and HLA-DR+ contribute to the IRS (10, 13, 14) by producing pro-inflammatory cytokines, including IL-2, IL-17 and IFN-γ (6, 7, 10, 13, 14). Activated Treg cells, including CD4+CD25+FoxP3+ cells that can express the cannabinoid receptor CB1, CD152 (CTLA-4) or glycoprotein A repetitions predominant (GARP) (7, 10, 13-15), promote CIRS by regulating the immune response and/or generating anti-inflammatory cytokines such as IL-10 and TGF-β (16). By modulating the production and secretion of pro-inflammatory cytokines (17), Treg cells play a crucial role in preserving immune balance and promoting immune tolerance. In animal studies, Treg depletion may amplify immune-inflammatory pathways and result in autoimmunity (18).

Using flow cytometry results, early precision medicine studies demonstrated that major depression, particularly melancholia, is qualitatively distinct from controls and minor depressed patients (19). The most significant CD markers were elevated surface Ig, CD25+, and HLA-DR+ expression on CD4+ T cells, indicating T cell activation in depression and particularly melancholia (19). In a separate study, Maes et al. (20) determined that 64% of patients with MDD had increased expression of CD7+CD25+ and CD2+HLADR+ cells with a specificity of 91%. According to these findings, numerous patients with MDD exhibit T cell activation. The soluble transferrin receptor (sTfR or sCD71) and sIL-2R (CD25+) are elevated in the serum of patients with MDD and BD (21-23), corroborating these findings.

Through their involvement in modulating cytokine production, Treg cells have been found to potentially impact mood disorders. Individuals with MDD have lower Treg levels than healthy individuals (17), and antidepressant-treated MDD patients have a greater number of CD4+CD25+ and CD4+CD25+FOXP3+Treg cells (24). The remitted phase of BD is characterized by a suppression of Teff and an activation of Treg, and increasing severity of BD, as measured by duration or number of manic episodes, is associated with Teff and Treg aberrations (10). Mice with depleted CD4+CD25+ Treg cells exhibit elevated levels of despair behaviors and decreased 5-HT in the hippocampus (25).

Nevertheless, no studies have examined whether activation of T cells and depletion of Treg cells are associated with increased severity of depression in an acute phase of MDD and whether MDD is characterized by very early (CD69+), early (CD71+), late (HLADR+) activation markers, and/or increases in CD40L+, a key player in T-cell dependent effector functions and humoral immunity (26-29). Consequently, the purpose of this study is to quantify the levels of CD69, CD71, CD40L an HLADR-bearing CD3 (pan T), CD4 and CD8 cells, and CB1, CD152 and GARP-bearing CD25+FoxP3+ Treg cells in the acute phase of MDD, and to determine if activation of T cells and depletion of Treg are associated with the severity of the acute phase. In addition, the relationships between these T cell subsets and immune cell profiles, based on cytokine production, are assessed.

## Methodology

### Participants

The investigation included participants from Bangkok’s Chulalongkorn University. The Department of Psychiatry’s senior psychiatrists recruited outpatients with MDD. Posters and word-of-mouth were used to recruit healthy controls. Before taking part in the study, all participants were required to sign a written consent form. Once participants were recruited, they were required to complete standard questionnaires and provide blood samples. Standard questionnaires and a semi-structured interview were utilized to collect demographic data, including age, gender, and body mass index. Patients between the ages of 18 and 65 who comprehended Thai, had been diagnosed with MDD by a psychiatrist using DSM-5 criteria, and had a Hamilton Depression Rating Scale (HAM-D) score greater than 17, indicating moderate to severe depression, met the inclusion criteria. Patients with other DSM-5 axis 1 disorders, including schizophrenia, schizoaffective disorders, obsessive compulsive disorder, post-traumatic stress disorder, psycho-organic disorders, or substance misuse disorders, were excluded from the study. Also precluded were healthy volunteers with a diagnosis of any axis 1 DSM-5 disorder or a positive family history of MDD, BD and psychosis. Patients and controls were ineligible if they had experienced any allergic or inflammatory responses in the previous three months, if they had neuroinflammatory, neurodegenerative, or neurological disorders such as epilepsy, Alzheimer’s disease, multiple sclerosis, or Parkinson’s disease, if they had (auto)immune diseases such as COVID-19 infection (lifetime), chronic obstructive pulmonary disease, inflammatory bowel disease, psoriasis, diabetes type 1, asthma, or rheumatoid arthritis, had a history of receiving immunomodulatory drugs, had taken therapeutic doses of antioxidants or omega-3 polyunsaturated fatty acid supplements within three months before the study, had used anti-inflammatory drugs (NSAID or steroids) within one month of the study. We did not include lactating or expectant women. Some patients were taking psychotropic medications, such as sertraline (18 patients), other antidepressants (8 patients, such as fluoxetine, venlafaxine, escitalopram, bupropion, and mirtazapine), benzodiazepines (22 patients), atypical antipsychotics (14 patients), and mood stabilizers (4 patients). In the statistical analysis, the potential effects of these drug variables were taken into consideration.

Before participating in the study, all controls and patients gave their written informed consent. The study was carried out following both international and Thai ethics and privacy regulations. The Institutional Review Board of the Faculty of Medicine at Chulalongkorn University in Bangkok, Thailand approved the study (#528/63), which was in accordance with the international guidelines for human research protection, such as the Declaration of Helsinki, the Belmont Report, CIOMS Guideline, and the International Conference on Harmonization in Good Clinical Practice (ICH-GCP).

### Measurements

A research assistant conducted a semi-structured interview and an experienced psychiatrist administered the 17-item version of the HAM-D to assess the intensity of depressive symptoms (30). The Mini-International Neuropsychiatric Interview (M.I.N.I.) was used to make the diagnosis of psychiatric disorders (31).

After an overnight fast, between 8:00 and 9:00 a.m., participants’ blood (20 mL) was collected. The blood was collected in BD Vacutainer® EDTA (10 mL) and BD Vacutainer® SST™ (5 mL) tubes provided by BD Biosciences (Franklin Lakes, NJ, USA). The serum-separating tubes were left to clot at room temperature for 30 minutes to obtain serum. The tubes were spun at 1100 ×g for 10 minutes at 4°C. Peripheral blood mononuclear cells (PBMCs) were separated from the blood sample through density gradient centrifugation (30 min at 900 ×g) using Ficoll® Paque Plus (GE Healthcare Life Sciences, Pittsburgh, PA, USA). Utilizing a hemocytometer and trypan blue, 0.4% solution, pH 7.2-7.3 (Sigma-Aldrich Corporation, St. Louis, Missouri, United States), the cell count and viability were determined. Counting total and blue-stained cells ensured that greater than 95% of cells were viable under all conditions. To activate PBMCs, the 96-well plates were coated with 5 µg/mL of the anti-human CD3 antibody (OKT3, from eBioscience), overnight. 3 x 10^5^ PBMCs were added to each well along with 5 μg/mL of the anti-human CD28 antibody (CD28.2, eBioscience). The cells were cultured in RPMI 1640 medium with L-glutamine supplemented with 10% fetal bovine serum and 1% penicillin-streptomycin (Gibco Life Technologies, Carlsbad, CA, USA) for 3 days at 37°C in an incubator with 5% CO2. The unstimulated PBMCs cultured for the same period were used as a negative control. After 3 days, the lymphocyte immunophenotypes were determined through flow cytometry.

To study lymphocyte immunophenotypes, 3×10^5^ PBMCs were labeled to surface markers with monoclonal antibodies for 30 minutes, including CD3-PEcy7, CD4-APCcy7, CD8-APC, CD40L-FITC, CD69-AF700, CD71-PerCPcy5.5, HLA-DR-PE594, CD25-APC, CD152 PE Dazzle594, GARP-PE, CB1-AF700 and 7-AAD (Biolegend, BD Biosciences and R&D Systems). For Treg cells, we firstly stained cells with surface markers (CD3, CD4, CD25) followed by intracellular FoxP3 staining, which was performed using the FoxP3/Transcription Factor Staining Buffer Set (eBioscience) for fixation and permeabilization before staining with antibody to Foxp3-FITC (Biolegend). Electronic Supplementary File (ESF1), Figures 1-2 show our gating strategies. The flow cytometry was performed using LSRII flow cytometer (BD Biosciences) to evaluate lymphocyte immunophenotypes. All data were analyzed using FlowJo X software (Tree Star Inc., Ashland, OR, USA). Table 1 of the ESF2 shows the different CD and cell surface markers, as well as their key functions, measured in the present study. We assessed both the percentages of T cells and the median fluorescence intensity (MFI) of the markers.

**Table 1.** Demographic and clinical data of the depressed patients and healthy controls (HC) included in the present study.

| **Variables** | **HC (n=20)** | **MDD (n=30)** | **F/W/$\chi^2$** | **df** | **p** |
| --- | --- | --- | --- | --- | --- |
| Sex (Male/Female) | 6/14 | 11/19 | $\chi^2=0.24$ | 1 | 0.626 |
| Age (years) | 33.6 (8.0) | 28.7 (8.6) | F=4.28 | 1/48 | 0.044 |
| Education (years) | 16.1 (2.2) | 15.6 (1.4) | F=0.84 | 1/48 | 0.363 |
| Smoking | 18/2 | 23/7 | FEPT | - | 0.285 |
| BMI (kg/m<sup>2</sup>) | 21.3 (2.5) | 25.5 (5.9) | F=8.82 | 1/48 | 0.005 |
| HAMD | 1.0 (1.6) | 23.5 (5.8) | F=289.87 | 1/48 | <0.001 |
| M1 macrophage (z scores) | -0.131 (0.116) | 0.087 (0.205) | W=0.86 | 1/48 | 0.354 |
| T helper-1 (z scores) | -0.337 (0.119) | 0.225 (0.198) | W=5.59 | 1/48 | 0.018 |
| IRS (z scores) | -0.366 (0.114) | 0.244 (0.184) | W=7.91 | 1/48 | 0.005 |
| T cell growth (z scores) | -0.305 (0.110) | 0.203 (0.169) | W=6.64 | 1/48 | 0.011 |
| Neurotoxicity (z scores) | -0.286 (0.125) | 0.190 (0.197) | W=4.09 | 1/48 | 0.043 |

Results are shown as mean ( ± SD). FEPT: Fisher's exact probability test; $X^2$ : analysis of contingency tables, F: results of analysis of variance, W: results of General estimating equation (GEE), the p values are results of pairwise comparisons between MDD and controls; BMI: body mass index; HAMD: Hamilton Depression Rating Scale score; IRS: immune-inflammatory responses system.

As previously described (15), we measured cytokines/chemokines/growth factors in unstimulated and stimulated diluted whole blood culture supernatant using the same blood samples as those employed for the flow cytometry. We used RPMI-1640 medium supplemented with L-glutamine, phenol red, and 1% penicillin (Gibco Life Technologies, USA), with or without 5 µg/mL PHA (Merck, Germany) + 25 µg/mL lipopolysaccharide (LPS; Merck, Germany). On sterile 24-well plates, 1.8 mL of each medium was combined with 0.2 mL of 1/10-diluted whole blood. Using the Bio-Plex Pro human cytokine 27-plex assay kit (BioRad, Carlsbad, California, United States of America) and the LUMINEX 200 instrument (BioRad, Carlsbad, California, United States of America), the cytokines/chemokines/growth factors were measured. The intra-assay CV values were less than 11%. ESF2, Table 2 shows the cytokines/chemokines/growth factors that were measured, and ESF2, Table 3 displays the immune profiles utilized in the current study.

**Table 2.** Differences in unstimulated (UNST) and stimulated (STIM) changes in the percentage of T lymphocyte populations in healthy controls (HC) and major depressed patients (MDD)

| **Variables**<br>**(z scores)** | **Baseline vs**<br>**anti-**<br>**CD3/CD28** | **HC**<br>**n=20** | **MDD**<br>**n=30** | **Pairwise**<br>**comparisons** |
| --- | --- | --- | --- | --- |
| CD3+CD69+% | UNST | -0.487 (0.090) | -0.479 (0.095) | 0.947 |
|  | STIM | 0.904 (0.024) | 0.921 (0.027) | 0.639 |
| **CD3+CD71+%** | **UNST** | **-1.673 (0.188)** | **-0.930 (0.142)** | **0.002** |
|  | STIM | 1.054(0.016) | 1.069 (0.017) | 0.520 |
| **CD3+CD40L+%** | **UNST** | **-0.955 (0.103)** | **-0.644 (0.059)** | **0.009** |
|  | STIM | 0.555 (0.026) | 0.553 (0.031) | 0.954 |
| CD3+HLADR+% | UNST | -0.229 (0.081) | -0.132 (0.065) | 0.350 |
|  | STIM | 0.730 (0.028) | 0.725 (0.021) | 0.510 |
| CD4+CD69+% | UNST | -0.571 (0.125) | -0.445 (0.107) | 0.444 |
|  | STIM | 0.907 (0.026) | 0.960 (0.024) | 0.130 |
| **CD4+CD71+%** | **UNST** | **-1.586 (0.187)** | **-0.802 (0.139)** | **0.001** |
|  | STIM | 1.099 (0.014) | 1.108 (0.014) | 0.636 |
| **CD4+CD40L+%** | **UNST** | **-0.818 (0.105)** | **-0.520 (0.059)** | **0.013** |
|  | STIM | 0.718 (0.024) | 0.694 (0.028) | 0.505 |
| **CD4+HLA-DR+%** | **UNST** | **-0.325 (0.071)** | **-0.113 (0.061)** | **0.024** |
|  | STIM | 0.676 (0.031) | 0.707 (0.022) | 0.411 |
| CD8+CD69+% | UNST | -0.748 (0.153) | -0.869 (0.126) | 0.541 |
|  | STIM | 0.831 (0.035) | 0.776 (0.044) | 0.319 |
| CD8+CD71+% | UNST | -1.982 (0.225) | -1.575 (0.174) | 0.152 |
|  | STIM | 1.060 (0.022) | 1.099 (0.018) | 0.139 |
| **CD8+CD40L+%** | **UNST** | **-2.026 (0.146)** | **-1.643 (0.096)** | **0.028** |
|  | **STIM** | **0.062 (0.055)** | **0.218 (0.056)** | **0.046** |
| CD8+HLADR+% | UNST | -0.369 (0.113) | -0.346 (0.083) | 0.869 |
|  | STIM | 0.793 (0.031) | 0.791 (0.0226) | 0.953 |

Results are shown as estimated marginal mean values (SE).

**Table 3.** Differences in unstimulated (UNST) and stimulated (STIM) changes in the percentage of T regulatory (Treg) lymphocyte populations in healthy controls (HC) and major depressed patients (MDD)

|  | Condition | Diagnosis |  | Tests of Model effects |  |  |
| --- | --- | --- | --- | --- | --- | --- |
| Variables<br>(z scores) |  | HC<br>n=20 | MDD<br>n=30 | Effects | Wald<br>df=1 | p |
| CD25+ FoxP3+CB1+% | UNST | **-0.238 (0.290)** | **-0.879 (0.123)** | G | 2.66 | 0.103 |
|  | STIM | 0.599 (0.124) | 0.643 (0.070) | G X T | 4.468 | **0.035** |
| CD25+ FoxP3+CD152+% | UNST | -0.390 (0.222) | -0.873 (0.159) | G | 3.565 | 0.059 |
|  | STIM | 0.780 (0.095) | 0.619 (0.077) | G X T | 1.659 | 0.198 |
| CD25+ FoxP3+GARP+% | UNST | -0.523 (0.216) | -0.907 (0.138) | G | 1.134 | 0.287 |
|  | STIM | 0.719 (0.079) | 0.778 (0.064) | G X T | 3.359 | 0.067 |

Results are shown as estimated marginal mean values (SE).

#### Statistics

Among study groups, we compared nominal variables via analysis of contingency tables (χ2-test) and continuous variables via analysis of covariance (ANCOVA). We employed Pearson’s product moment correlation coefficients for the purpose of examining the relationships between scale variables. A pre-specified generalized estimating equation (GEE), repeated measures, included fixed categorical effects of diagnosis (differences between patients and controls), cell types (T cell populations, namely CD69, CD71, CD40L, and HLADR-bearing CD3+, CD4+ and CD8+ cells), and the responsivity of the cells to in vitro administration of anti-CD3/CD28 beads (unstimulated versus stimulated condition), while allowing for the effects of age, sex, the drug state, BMI and smoking. Using pairwise comparisons, predefined comparisons were examined. Prespecified GEE, repeated measures, were used to examine the fixed categorical effects of diagnosis, treatment (unstimulated versus stimulated), cell type, and their interactions on Treg cells (CD25+FoxP3+CB1+, CD25+FoxP3+CD152+, and CD25+FoxP3+GARP+), while allowing for the effects of the aforementioned background variables. The primary statistical analyses examined the effects of Teff and Treg subsets on the HAMD score using multiple regression analyses. These analyses are the most important because a) a quantitative outcome score such as the HAMD is much more informative or correct than the binary diagnosis MDD (Maes et al., 2022; Maes and Almulla, 2023), and b) this method allows to examine the combined effects of both activated T and Treg cells on severity of illness. Using manual multiple regression analysis, the effects of cell populations on the HAMD score were evaluated. We utilized an automatic forward stepwise regression strategy with a p-to-enter of 0.05 and a p-to-remove of 0.06 to determine which variables would be included and which would be excluded from the final regression model. We estimated the standardized β coefficients with t-statistics and exact p-values for each explanatory variable, F-statistics (and p-values), and partial eta squared (effect size) of the model. Using the modified Breusch-Pagan test and the White test, heteroskedasticity was investigated. We examined the probability of multicollinearity and collinearity utilizing the tolerance (cut-off value: 0.25) and variance inflation factor (cut-off value: > 4), as well as the condition index and variance proportions from the collinearity diagnostics table. Residuals, residual plots, and data quality were always evaluated in the final model. We also computed partial regression analyses, including partial regression plots, based on the results of the linear modeling analyses. When required, we utilized transformations such as Log10, square-root, rank-based inversed normal, and Winsorization to normalize the data distribution of the indicators. At p=0.05, all analyses were two-tailed. To conduct the aforementioned statistical analyses, we utilized IBM’s Windows version of SPSS 28. Given an effect size of 0.25 (equivalent to 20% explained variance), alpha=0.05, power=0.8, and 3 covariates, an a priori power analysis (G*Power 3.1.9.4) for a linear multiple regression suggests that the minimum sample size should be 48.

## Results

### Demographic data

**Table 1** displays the sociodemographic and clinical characteristics of the study’s patients and controls. There were no significant differences between the study groups in terms of age, gender distribution, education, or smoking. Patients had a higher BMI than controls. In any case, we have controlled all results for possible effects of age, BMI, sex, smoking, and the drug state of the patients but could not find any significant effects. The average HAMD scores of patients were substantially higher than those of controls, indicating that the majority of patients suffered from moderate to severe clinical depression. The outcomes of GEE analyses with immune profiles as dependent variables and stimulation status (baseline vs LPS+PHA treatment), diagnosis (MDD versus controls), and stimulation by diagnosis interaction as predefined fixed effects are presented in the same table. The Th-1, IRS, T cell growth, and neurotoxicity (but not M1) profiles were all higher in MDD individuals compared to controls.

### Baseline and stimulated Teff frequencies and MFI in MDD

GEE analysis performed on the frequency of activated T cells showed significant effects of diagnosis (W=4.78, df=1, p=0.029), stimulation (Wald=596.28, df=1, p<0.001)), cell type (W=1121.92, df=1, p<0.001)), diagnosis X cell type (W=30.59, df=11, p=0.001), stimulation X cell type (W=542.25, df=1, p<0.001), and the three-way interaction cell type X diagnosis X stimulation (W=25.72, df=11. P=0.007). There were no significant effects of age, sex, BMI, smoking or drug state of the patients. **Table 2** shows the frequencies of the T cell subtypes according to diagnosis and stimulation status. Anti-CD3/CD28 stimulation significantly increased the frequencies of all subsets. Activation markers were significantly higher in MDD (29.34 ±0.98 %) than in controls (27.6±0.93 %). ESF2, Table 4 shows the measurements of the 4 activation markers in CD3+, CD4+ and CD8+ T cells in baseline and stimulated conditions in MDD patients and controls. Pairwise comparisons showed that the number of activated T cells in the baseline condition was greater in patients than controls (p=0.041). Table 2 shows that in the baseline condition, MDD patients showed higher frequencies of baseline CD3+CD71+, CD3+CD40L+, CD4+CD71+, CD4+CD40L+, CD4+HLA-DR+ and CD8+CD40L+ than normal controls. After stimulation with anti CD3/CD28 beads, there were no significant differences in any of the cell types between MDD and controls, except CD8+CD40L+ which was elevated in MDD.

**Table 4.** Results of multiple regression analyses with the Hamilton Depression rating Scale (HAMD) score as a dependent variable

| Dependent Variables | Explanatory Variables | Parameter estimates |  |  | Model |  |  |  |
| --- | --- | --- | --- | --- | --- | --- | --- | --- |
| | | $\beta$ | t | p | F | df | P | R<sup>2</sup> |
| #1. HAMD | Model |  |  |  | 8.94 | 3/44 | <0.001 | 0.379 |
|  | U_CD25+FOXP3+CB1+ % | -0.460 | -3.77 | <0.001 |  |  |  |  |
|  | S_CD4+CD69+ % | 0.400 | 3.28 | 0.002 |  |  |  |  |
|  | U_CD4+CD40L+ % | 0.284 | 2.38 | 0.022 |  |  |  |  |
| #2. HAMD | Model |  |  |  | 5.85 | 2/45 | 0.006 | 0.206 |
|  | Age | -0.344 | -2.59 | 0.013 |  |  |  |  |
|  | S_CD8+CD40L+ % | 0.311 | 2.34 | 0.024 |  |  |  |  |
| #3. HAMD | Model |  |  |  | 5.91 | 4/43 | <0.001 | 0.355 |
|  | Age | -0.401 | -3.13 | 0.003 |  |  |  |  |
|  | S_CD8+CD40L+ MFI | 0.341 | 2.53 | 0.015 |  |  |  |  |
|  | S_CD3+HLADR+ MFI | 0.362 | 2.65 | 0.011 |  |  |  |  |
|  | S_CD4+CD69+ MFI | 0.270 | 2.04 | 0.048 |  |  |  |  |
| #4. HAMD | Model |  |  |  | 6.21 | 4/43 | <0.001 | 0.366 |
|  | Age | -0.417 | -3.29 | 0.002 |  |  |  |  |
|  | S_CD8+CD40L+ MFI | 0.426 | 3.31 | 0.002 |  |  |  |  |
|  | S_CD3+HLADR+ MFI | 0.373 | 2.75 | 0.009 |  |  |  |  |
|  | U_CD25+FOXP3+CB1+ MFI | -0.278 | -2.24 | 0.030 |  |  |  |  |
| #5. HAMD | Model |  |  |  | 4.32 | 1/44 | 0.044 | 0.089 |
|  | U_CD25+FOXP3+GARP+ MFI | -0.299 | -2.99 | 0.044 |  |  |  |  |

%; prevalence on immune cell populations; MFI: median fluorescence intensity; S: stimulated, U: unstimulated.

ESF2, Table 5 shows the outcome of a similar GEE analysis performed on the MFI values of the activated T cells. This GEE analysis showed significant effects of anti-CD3/CD28 stimulation (Wald=825.31, df=1, p<0.001), cell type (W=1875.74, df=1, p<0.001), diagnosis X cell type (W=23.00, df=11, p=0.018), stimulation X cell type (W=1133.54, df=1, p<0.001), and the three-way interaction cell type X diagnosis X stimulation (W=24.12, df=11. P=0.012). ESF2, Table 5 displays that MDD patients exhibit increased baseline MFI values of CD3+CD71+, CD4+CD71+, and CD4+HLADR+ and a trend toward increased CD3+HLADR+ MFI values as compared with controls. In addition, pairwise comparisons showed significant increases in stimulated CD3+CD71+ and CD4+HLADR+ MFI values between MDD patients and controls (p<0.05). There were no significant effects of age, sex, BMI, smoking or drug state of the patients.

**Table 5.** Results of multiple regression analyses with immune profiles as dependent variables and T cell subsets or expression markers as explanatory variables.

| Dependent Variables | Explanatory Variables | Estimates |  |  | Model |  |  |  |
| --- | --- | --- | --- | --- | --- | --- | --- | --- |
| | | $\beta$ | t | p | F | df | p | R<sup>2</sup> |
| #1. M1 | Model |  |  |  | 6.48 | 2/45 | 0.003 | 0.224 |
|  | S_CD25+FoxP+CD152+ % | -0.391 | -2.98 | 0.005 |  |  |  |  |
|  | S_CD8+CD69+ % | 0.271 | 2.06 | 0.045 |  |  |  |  |
| #2. Th-1 | Model |  |  |  | 5.13 | 1/46 | 0.030 | 0.099 |
|  | S_CD25+FoxP3+CD152+ % | -0.314 | -2.24 | 0.030 |  |  |  |  |
| #3. IRS | Model |  |  |  | 5.92 | 3/44 | 0.002 | 0.288 |
|  | S_CD4+CD69+ % | 0.338 | 2.62 | 0.012 |  |  |  |  |
|  | U_CD25+FoxP3+CD152+ % | -0.336 | -2.57 | 0.014 |  |  |  |  |
|  | S_CD4+CD71+ % | 0.287 | 2.16 | 0.036 |  |  |  |  |
| #4. T cell growth | Model |  |  |  | 8.03 | 3/44 | <0.001 | 0.354 |
|  | S_CD4+CD69+ % | 0.378 | 3.07 | 0.004 |  |  |  |  |
|  | U_CD25+FoxP3+CD152+ % | -0.366 | -2.96 | 0.005 |  |  |  |  |
|  | U_CD3+CD71+ % | 0.301 | 2.40 | 0.021 |  |  |  |  |
| #5. Neurotoxicity | Model |  |  |  | 4.99 | 1/46 | 0.030 | 0.098 |
|  | S_CD25+FoxP3+GARP+ % | -0.313 | -2.23 | 0.030 |  |  |  |  |
| #6. M1 | Model |  |  |  | 7.12 | 2/45 | 0.002 | 0.240 |
|  | S_CD25+FoxP3+CD152+ MFI | -0.398 | -3.04 | 0.004 |  |  |  |  |
|  | S_CD8+CD69+ MFI | 0.346 | 2.64 | 0.011 |  |  |  |  |
| #7. Th-1 | Model |  |  |  | 6.81 | 1/46 | 0.012 | 0.129 |
|  | S_CD8+CD71+ MFI | 0.359 | 2.61 | 0.012 |  |  |  |  |
| #8. IRS | Model |  |  |  | 10.10 | 1/46 | 0.003 | 0.180 |
|  | S_CD8+CD71+ MFI | 0.424 | 3.18 | 0.003 |  |  |  |  |
| #9. T cell growth | Model |  |  |  | 7.47 | 3/44 | <0.001 | 0.338 |
|  | S_CD3+CD71+ MFI | 0.354 | 3.54 | <0.001 |  |  |  |  |
|  | S_CD4+CD69+ MFI | 0.325 | 2.62 | 0.012 |  |  |  |  |
|  | S_CD25+FoxP3+CD152+ MFI | -0.299 | -2.43 | 0.019 |  |  |  |  |
| #10.<br>Neurotoxicity | Model |  |  |  | 6.95 | 2/43 | 0.002 | 0.244 |
|  | U_CD25+FoxP3+ CD152+ MFI | -0.489 | -3.48 | 0.001 |  |  |  |  |
|  | U_CD8+CD40L+ MFI | 0.339 | -2.41 | 0.020 |  |  |  |  |

%; prevalence on immune cell populations; MFI: median fluorescence intensity; S: stimulated, U: unstimulated.

GEE analysis performed on the frequencies of the three CD25+FoxP3+ subtypes showed a significant effect of diagnosis (W=3.96, df=1, p=0.047), stimulation (W=211.51, df=1, p<0.001), cell type (W=529.81, df=1, p<0.001) and cell type X simulation (W=243.14, df=1, p<0.001). The CD25+FoxP3+ cells were significantly lower in MDD (z score ±SE: -0.073 ±0.055) than in controls (0.111 ±0.074). As can be seen in **Table 3**, in vitro administration of anti-CD3/CD28 beads increased the number of all three FoxP3 cell types significantly. There was a significant group by time interaction for CD25+FoxP3+CB1, indicating that patients with MDD showed significantly lower CD25+FoxP3+CB1+ levels than controls. The results of GEE, repeated measures, performed on the MFI of the CD25+FoxP+ cells shows a significant effect of stimulation (W=75.58, df=1, p<0.001) and cell types (W=39.52, df=1, p<0.001). ESF2, Table 6 shows the interaction between diagnosis and stimulation with anti-CD3/CD28. Overall, stimulation significantly increased the MFI of FoxP3 cells. Pairwise comparisons show that the unstimulated expression of FoxP3 was significantly lower in MDD as compared with controls (p=0.027), whereas there were no significant intergroup differences in the stimulated condition (p=0.772).

**Table 6.** Results of general equating estimation, repeated measurement

| **Dependent Variables** | **Explanatory Variables** | ***B*** | **SE** | **Lower** | **Upper** | **Wald** | **df** | ***p*** |
| --- | --- | --- | --- | --- | --- | --- | --- | --- |
| IRS | U_CD25+FoxP3+CD152+ % | -0.233 | 0.0588 | -0.348 | -0.117 | 15.65 | 1 | <0.001 |
|  | S_CD4+CD71+ % | 1.190 | 0.0775 | 1.638 | 1.342 | 235.98 | 1 | <0.001 |
| IRS | U_CD25+FoxP3+CD152+ % | -0.185 | 0.0467 | -0.277 | -0.094 | 15.80 | 1 | <0.001 |
|  | U_CD8+CD40L+ % | 0.520 | 0.1948 | 0.138 | 0.902 | 7.12 | 1 | 0.008 |
| IRS | S_CD8+CD40L+ MFI | 3.199 | 0.1649 | 2.876 | 3.522 | 376.43 | 1 | <0.001 |
|  | U_CD25+FoxP3+CD152+ MFI | -0.197 | 0.0537 | -0.302 | -0.092 | 13.46 | 1 | <0.001 |
| Neurotoxicity | U_CD25+FoxP3+CD152+ % | -0.158 | 0.0541 | -0.258 | -0.058 | 9.17 | 1 | 0.002 |
|  | CD8+CD40L+ % | 1.958 | 0.0906 | 1.780 | 2.135 | 466.52 | 1 | <0.001 |
| Neurotoxicity | CD8+CD40L+ MFI | 3.420 | 0.1488 | 3.129 | 3.712 | 588.62 | 1 | <0.001 |
|  | U_CD25+FoxP3+CD152+ MFI | -0.174 | 0.0479 | -0.268 | -0.085 | 13.13 | 1 | <0.001 |

IRS: immune-inflammatory response system; U: unstimulated; S: stimulated; MFI: median fluorescence intensity

### Regression analyses of severity of illness on T cell subtype percentages and MFI values

Consequently, we performed the primary outcome statistical analyses, namely multiple regression analyses with the HAMD as outcome variable and the different cell populations (either frequencies or MFI) as explanatory variables. **Table 4**, regressions #1 and #2 show the outcome of these regressions using the percentages of the cell types, whereas regressions #4, #5 and #6 show the outcome of regressions on the MFI values. We examined the effects of the unstimulated and stimulated cell types. Regression #1 shows that 37.9% of the variance in the HAMD could be explained by unstimulated CD25+FoxP3+CB1 % (inversely), unstimulated CD4+CD40L+ % and stimulated CD4+CD69+ % (positively). **Figure 1** shows the partial regression of the total HAMD score on the baseline number of CD4+CD40L+ cells. Nevertheless, also CD4+CD71+ % was significantly associated with the HAMD score as shown in **Figure 2**. Regression #2 (we omitted the CD4+ populations) shows part of the variance in the HAMD score could be explained by the regression on CD8+CD40+% (positively) and age (inversely).

Regarding the MFI data, we found that (regression #3) 35.5% of the variance in the HAMD could be explained by 3 stimulated MFI values (CD8+CD40L+, CD3+HLADR+, CD4+CD69+, all positively) and age (inversely). Regression #4 shows that this prediction could be slightly improved by adding the unstimulated CD25+FoxP3+CB1 MFI data. Regression #5 displays that unstimulated CD25+FoxP3+GARP+ MFI was inversely associated with the HAMD.

### Regression analyses of immune profiles on T cell subtype percentages and MFI values

**Table 5** shows the results of multiple regression analyses with immune profiles (M1, Th-1, IRS, T cell growth and neurotoxicity) as dependent variables and T cells, either percentage (regressions 1-5) or MFI (regressions 6-10) as explanatory variables, while allowing for the effects of extraneous variables. Regression #1 shows that 22.4% of the variance in M1 could be explained by CD8+CD69+ % (positively) and CD25+FoxP3+CD152+ % (inversely). Th-1 was best predicted by CD25+FoxP3+CD152+ % (inversely). Regression #3 shows that a larger part (28.8%) of the variance in IRS was explained by baseline CD25+FoxP3+CD152+ % (inversely) and stimulated CD4+CD69+ % and CD4+CD71+ % (positively). Regression #4 shows that the T cell growth profile was predicted by three different cell types, namely CD3+CD71+%, CD4+CD69+ % (positively) and CD25+FoxP3+CD152+ (inversely). Neurotoxicity was best predicted by CD25+FoxP3+GARP+ % (inversely).

With regard to the MFI values, we detected that M1 (regression #6) was best predicted by CD25+FoxP3+CD152+ MFI (inversely) and CD8+CD69+ MFI (positively). Regression #7 shows that 12.9% of the variance in Th-1 was predicted by CD4+CD71+ MFI (positively). IRS was strongly associated with CD8+CD71+ MFI (regression #8). T cell growth was predicted (33.8% of the variance) by CD3+CD71+ MFI, CD4+CD69+ MFI (positively), and CD25+FoxP3+CD152+ MFI (inversely). Regression #10 shows that 24.4% of the variance in neurotoxicity was explained by the unstimulated CD25+FoxP3+ CD152+ MFI (inversely) and CD8+CD40L+ MFI values.

**Table 6** shows that the changes from baseline to the LPS+PHA-stimulated IRS and neurotoxicity scores were predicted by lowered numbers or expression of CD25+FoxP3+CD152+ T reg cells combined with baseline or stimulated CD8+ and CD4+ T activation subsets (either prevalence or MFI values).

## Discussion

### Associations between T cells and immune profiles

The first major finding of this study is that there is a strong correlation between the flow cytometric assessments of T cell subtypes and cytokine-based immune-inflammatory profiles, including M1, Th-1, IRS, T cell growth, and neuroimmunotoxic profiles. Thus, these immune profiles were a) positively associated with the number or expression of diverse activated T cells, primarily CD4+CD71+ and CD8+CD69+ T cells, but also CD4+CD69+ T, CD8+CD71+ and CD8+CD40L+ cells, and b) negatively associated with the number or expression of diverse Treg cells, including CD152- and GARP-bearing CD25+FoxP3+ cells.

During T cell activation, distinct phases can be distinguished by the presence of distinct markers. CD69 is a cell surface marker that is expressed immediately after T cell activation and rapidly disappears after the initial stimulation (14). CD69 is a costimulatory molecule for T cell differentiation, activation, and proliferation (32) as well as lymphocyte retention in tissues (14).

CD69 activation stimulates the production of IL2 and TGF-β1 and controls the production of pro-inflammatory cytokines such as IL-17, IFN-γ, and IL-22, as well as Treg cells (14). CD40L is present in the early phases of T cell activation (33) and binds to CD40 on macrophages, CD8+ T cells, B cells, and dendritic cells, thereby promoting the differentiation and proliferation of immune cells, immune-inflammatory responses, and activation and maturation of B cells, which ultimately results in the production of autoantibodies (33). After antigen presentation, preformed CD40L molecules are mobilized from the lysosomes of Th-1 cells (34), while coligation of CD3 and CD40L increases the production of IFN-γ, TNF-α, and IL-10 (26). During a later phase of T cell activation, CD71 (TfR) is upregulated, resulting in an increase in iron uptake into the cells (35), which stimulates T cell activation and proliferation (36). Within twenty-four to forty-eight hours of T cell activation, the late phase activation marker HLA-DR is expressed. This latter molecule is a class II MHC, and its upregulation is associated with an increase in IFN-γ production (37).

Conversely, the inverse associations of the immune profiles with CD152 (CTLA-4) and GARP-bearing Treg cells indicate immunoregulatory Treg effects on different aspects of the immune response. Depending on the surface expression of the proteins, Tregs regulate the immune system through a variety of mechanisms. CTLA-4 or CD152 is expressed on Treg cells (and activated T cells) and functions as a negative immune checkpoint that regulates immune responses (38). By competing with CD28 for binding to the same ligands on antigen-presenting cells, CD152 inhibits the activation and proliferation of T cells (39). GARP is a Treg cell surface molecule that indicates Treg activation and the release of TGF-β1, an anti-inflammatory cytokine. As a result, Treg cells promote immune tolerance (39, 40). Treg cells, may express CB1, which inhibits the production of pro-inflammatory cytokines and exerts negative immunoregulatory effects (41). Previously, we discovered that the number of CD25+FoxP3+CB1+ cells was inversely associated with IRS responses (15). However, after introducing the effects of GARP- and CD152-bearing Tregs, the effects of CB1 were no longer significant. Intriguingly, the current study detected that decreased numbers of baseline Treg cells predict increased neurotoxic potential, indicating that diminished fitness of Treg cells is associated not only with exaggerated immune responses, but also with increased neuroimmunotoxicity.

### T cell activation in MDD

The second major finding of this study is that the number of baseline CD3+CD71+, CD3+CD40L+, CD4+CD71+, CD4+CD40L+, and CD4+HLADR+, expression of CD3+CD71+ CD+CD71+, CD4+HLADR+, and CD4+HLADR+ are significantly greater in MDD than in controls, and that the severity of depression is strongly predicted by increased CD4+CD40L+ and CD4+CD69+ numbers and the expression of CD40L on CD8+ cells and HLADR on CD3+ cells.

These results suggest that patients with MDD have an abnormal distribution of CD4+ and CD8+ T cell subsets with elevated T cell activation markers. Maes et al. demonstrated in previous investigations that depressed patients had a higher expression of T cell activation markers, specifically CD25+ and HLADR+ (20). A recent meta-analysis also reported an increase in the mean absolute number of activated T cells expressing CD25+ and CD3+HLADR+ (42). Moreover, Maes et al. (10) found that in the symptomatic remission phase of BD, the frequency of unstimulated CD3+ CD8+ CD71+ cells is lower than in healthy controls. This suggests that the acute phase of major mood disorders is characterized by T cell activation and that this T cell activation is suppressed when the symptoms ameliorate. Therefore, future research on T cell activation (and the immune system in general) should always differentiate between the acute, partially remitted, and remitted phases of mood disorders.

Our findings indicate that the pathogenesis of the acute phase of severe MDD is associated with CD4+ and CD8+ T cell activation. Interaction with the antigen-MHC complex activates CD4+ T cells, which consequently differentiate into distinct subtypes under the influence of cytokines (39). Almulla et al. (43) reported that increased IL-16, Th-1 activation and Th-1 polarization are hallmarks of the acute phase of severe MDD. IL-16 signaling occurs via the CD4 molecule on Th cells, activating CD4+ cells and upregulating activation markers and the IRS (44-46). Ligation of CD40 via CD40L stimulates the secretion of the Th-1-polarizing cytokine IL-12 (47, 48). Moreover, CD40L expression by CD4+ cells is a crucial step in the activation of CD8 T cells (29), which may express CDL40, perform T helper functions, and promote their own growth (27, 29). In addition, CD40L-CD40 interactions mediate the effect of CD4+ cells on CD8+ cytotoxic T lymphocytes (49)

Consequently, our findings suggest that CD4+ and CD8+ T cell activation, along with increased Th-1 polarization and IL-16 production, are key phenomena that drive increased T effector, T helper, and T cytotoxic functions, as well as enhanced M1, Th-17, IRS, and neurotoxic immune profiles in the acute phase of severe MDD. These activation markers may contribute to the immune, autoimmune, and neurotoxic processes seen in MDD. First, CD69 is involved in systemic lupus erythematosus (SLE), rheumatoid arthritis, atopic dermatitis, and systemic sclerosis, as well as animal models of arthritis, myocarditis, and inflammatory bowel disease (14). Second, increased expression of CD71 on T cells is associated with SLE disease activity and an enhanced Th-17 profile, whereas CD71 may cause autoimmunity in an SLE model (50). Targeting CD71 reduces autoimmunity and pathology in a mouse model of SLE and increases the secretion of IL-10 (50). Third, CD40L-CD40 interactions play a significant role in neurodegenerative and neurological disorders, such as Parkinson’s disease, Alzheimer’s disease, stroke, multiple sclerosis, and epilepsy (51). Increased permeability of the blood-brain barrier (BBB), injury to neuronal and glial cells, neuroinflammation, and the formation of microthrombi are mediated by CD40L-CD40 signaling (51). In an animal model of autoimmune disease, CD4+ immune cells, attracted and activated by IL-16, are associated with neuroinflammation (52). CD40L+ T cells may cross the BBB and activate microglial CD40, resulting in the production of pro-inflammatory cytokines and reactive nitrogen species, which may cause demyelination of axons (51). In addition, activated CD8+ cells may induce IRS responses and induce neurotoxicity (53).

### 3. Breakdown of immune tolerance in MDD

The third major finding of this study is that lowered numbers/expression of CD3+CD4+CD25+FoxP3+GARP+ and CD3+CD4+CD25+FOXP3+CB1+ coupled with T cell activation, is inversely associated with the severity of illness. Jahangard et al. showed that the frequency of FOXP3+ Tregs was decreased in untreated MDD patients compared to healthy controls, while the proliferation of circulating CD4+ T cells was increased in the patients (54). Ellul et al. (55) found an association between increased risk of MDD and decreased Treg numbers in association with increased inflammation (55). Importantly, during the remission phase of mood disorders, Treg functions may be upregulated (10).

Treg cells and CIRS activities ensure immune homeostasis by regulating innate and adaptive immune effector cells and restraining the detrimental activities of these effector cells. The transcription factor FoxP3 is a key regulatory factor in Treg activities by producing CIRS cytokines, such as IL-10 and TGF-β1, depleting growth factors, and expressing the co-inhibitory CTLA-4 molecule (56). Moreover, CD25+ (or IL-2R) Treg cells may absorb IL-2 precluding IL-2 for the growth of CD8+ T effector and Th-1 cells (57). Depletion of Treg cells may be the consequence of reduced fitness, altered apoptosis, and increased inhibitory activities of CD71 (50). Overall, the findings indicate that the acute phase of severe MDD is characterized by a breakdown of peripheral immune tolerance. Loss of Treg-associated immune tolerance can lead to immunopathologies and autoimmune diseases (58). In addition, Treg cells have neuroprotective properties by suppressing neurotoxic T effector responses and production of neurotoxic cytokines, and favoring anti-inflammatory microglia polarization (6, 57). When the frequency of Tregs is decreased or their immunosuppressive effects are diminished, the Teff/Treg balance is shifted towards the production of pro-inflammatory Th-1 and Th-7 phenotypes (57). As such, diminished Treg functions predispose to neuroinflammatory disorders including multiple sclerosis, stroke, and Alzheimer’s disease. These findings further support the IRS/CIRS theory of mood disorders and previous studies that have highlighted the imbalance of IRS and CIRS activity in patients with MDD (55, 59-61).

## Conclusions

The acute phase of severe MDD is characterized by a breakdown of immune tolerance, and CDL40 activation, which are associated with increased neuroimmunotoxic potential. Diminished Treg neuroprotection, T cell activation, Th-1 polarization, increased CD71+ and CD40L expression and IL-16 production are new drug targets to treat the T effector-associated neurotoxicity of the acute phase of severe MDD.

There is now some evidence that anti-inflammatory agents may be useful to treat MDD patients with an elevated immune profile (62). A recent meta-analysis shows that different types of anti-inflammatory agents have some clinical efficacy in treating MDD (63). Nevertheless, our results show that MDD should not be treated with repurposing anti-inflammatory drugs (e.g., Cox-2 inhibitors) and antioxidant supplements (e.g., curcumin), but rather should boost Treg functions and target CD40L or CD71 and IL-16 to attenuate T cell activation.

## Supporting information

ESF1

ESF2

## Data Availability Statement

The dataset generated during and/or analyzed during the current study will be available from the corresponding author (M.M.) upon reasonable request and once the dataset has been fully exploited by the authors

## Author’s contributions

Design of the study: MM and MR. Recruitment of the participants: MR and KJ. Assays: PS. Statistical analyses: MM. Visualization: MM and PS. Editing: MR, KJ, AS, and PS. All authors agreed to publish the final version of the manuscript.

## Ethical statement

All subjects gave their written informed consent for inclusion before they participated in the study. The study was conducted in accordance with the Declaration of Helsinki of 1975, revised in 2013, and the protocol was approved by the Ethics Committee of the Institutional Review Board of the Faculty of Medicine, Chulalongkorn University, Bangkok, Thailand (#528/63).

