## Supplementary material for "T cell activation via the CD40 ligand and transferrin receptor and deficits in T regulatory cells are associated with major depressive disorder and severity of depression": ESF1

### Slide 1
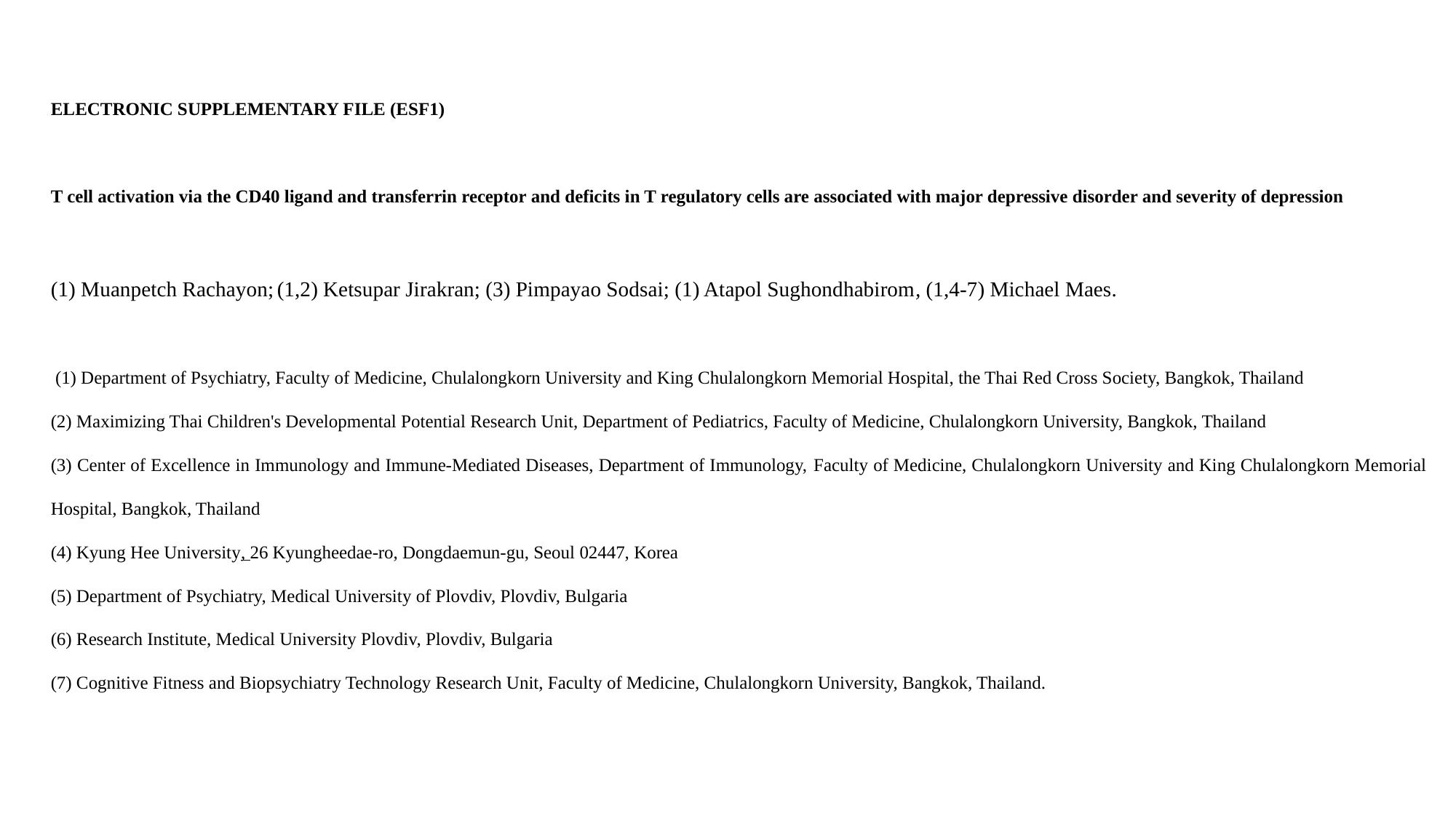

ELECTRONIC SUPPLEMENTARY FILE (ESF1)
T cell activation via the CD40 ligand and transferrin receptor and deficits in T regulatory cells are associated with major depressive disorder and severity of depression
(1) Muanpetch Rachayon; (1,2) Ketsupar Jirakran; (3) Pimpayao Sodsai; (1) Atapol Sughondhabirom, (1,4-7) Michael Maes.
 (1) Department of Psychiatry, Faculty of Medicine, Chulalongkorn University and King Chulalongkorn Memorial Hospital, the Thai Red Cross Society, Bangkok, Thailand
(2) Maximizing Thai Children's Developmental Potential Research Unit, Department of Pediatrics, Faculty of Medicine, Chulalongkorn University, Bangkok, Thailand
(3) Center of Excellence in Immunology and Immune-Mediated Diseases, Department of Immunology, Faculty of Medicine, Chulalongkorn University and King Chulalongkorn Memorial Hospital, Bangkok, Thailand
(4) Kyung Hee University, 26 Kyungheedae-ro, Dongdaemun-gu, Seoul 02447, Korea
(5) Department of Psychiatry, Medical University of Plovdiv, Plovdiv, Bulgaria
(6) Research Institute, Medical University Plovdiv, Plovdiv, Bulgaria
(7) Cognitive Fitness and Biopsychiatry Technology Research Unit, Faculty of Medicine, Chulalongkorn University, Bangkok, Thailand.

### Slide 2
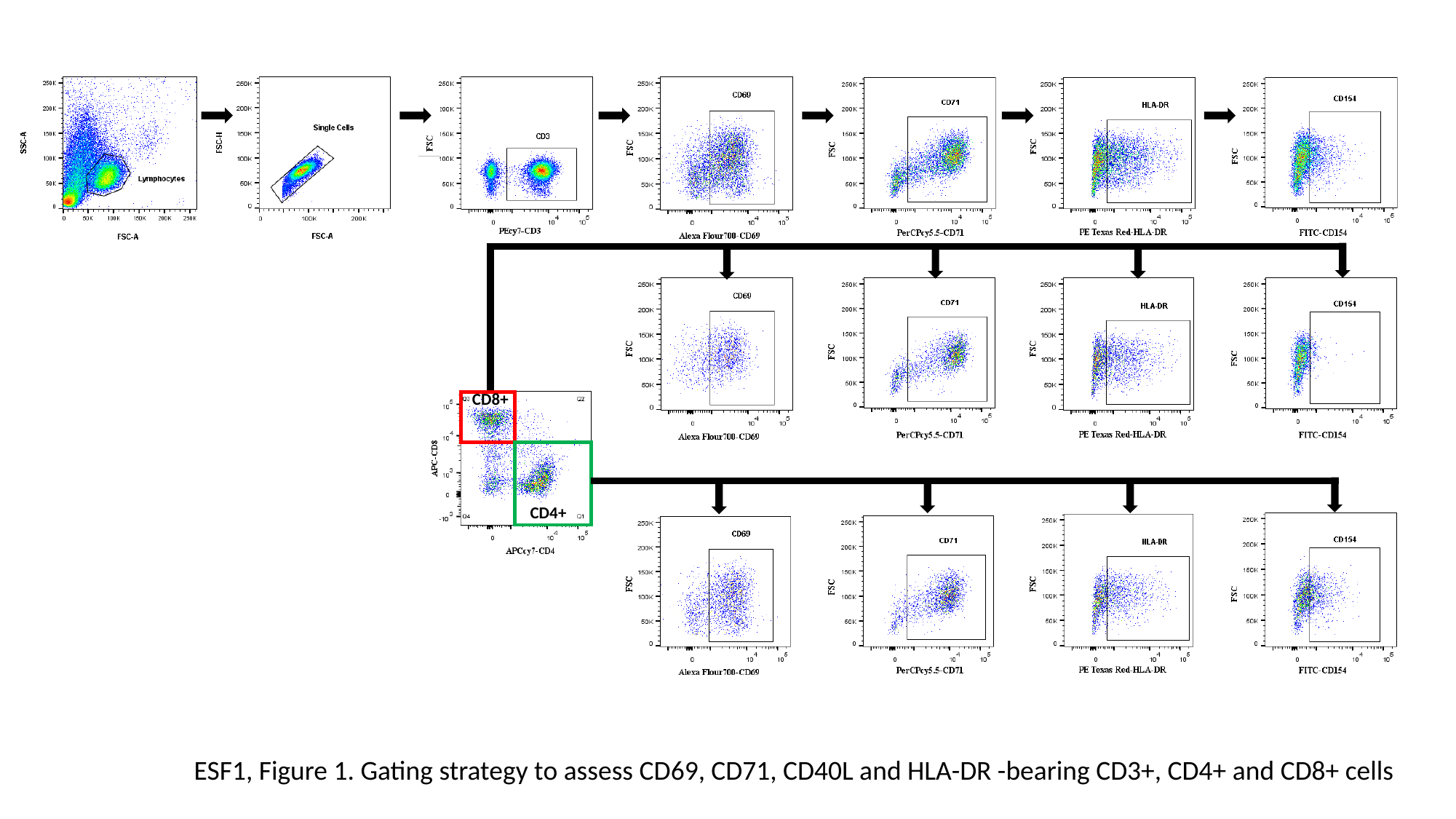

CD8+
CD4+
ESF1, Figure 1. Gating strategy to assess CD69, CD71, CD40L and HLA-DR -bearing CD3+, CD4+ and CD8+ cells

### Slide 3
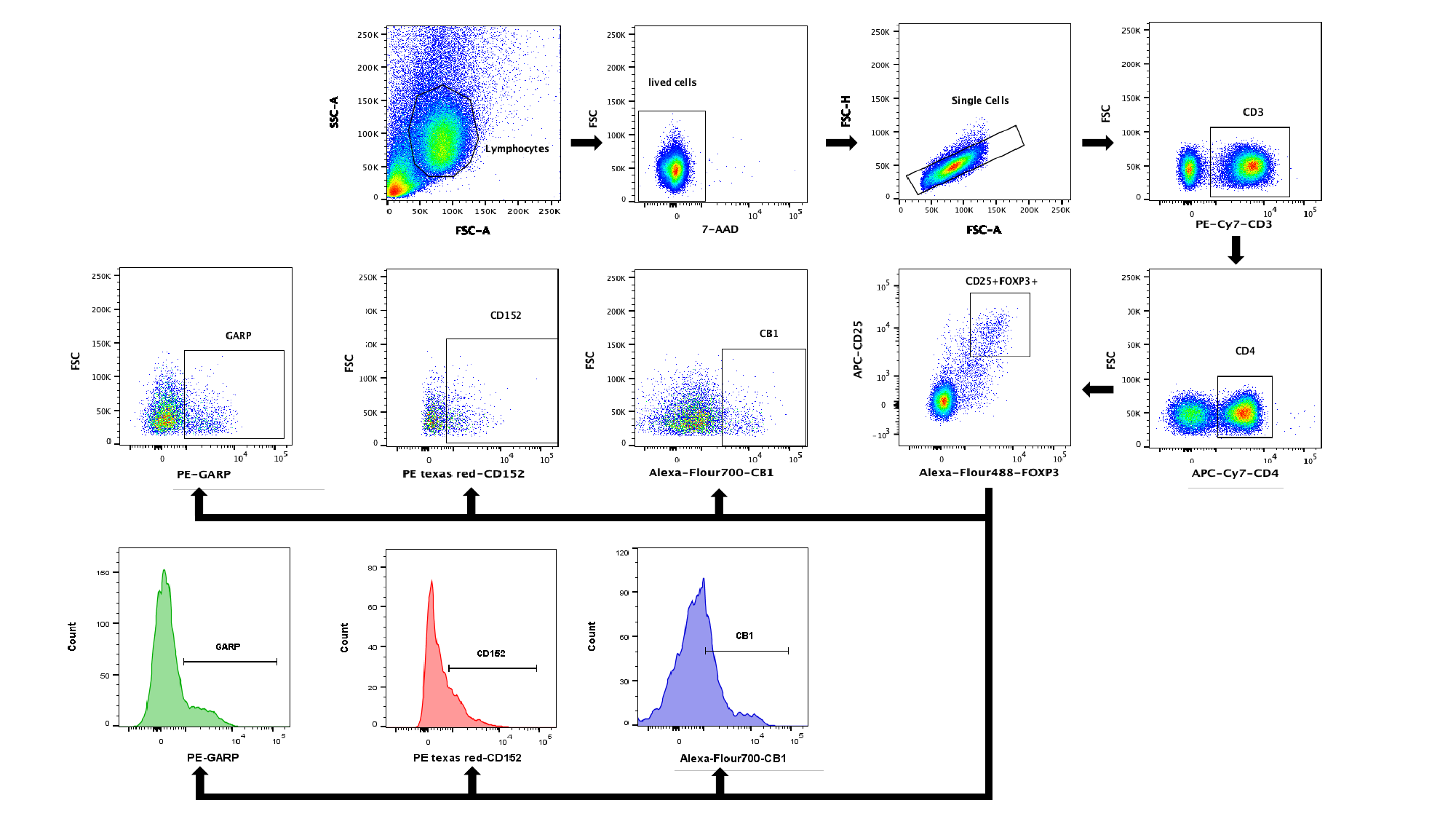

### Slide 4
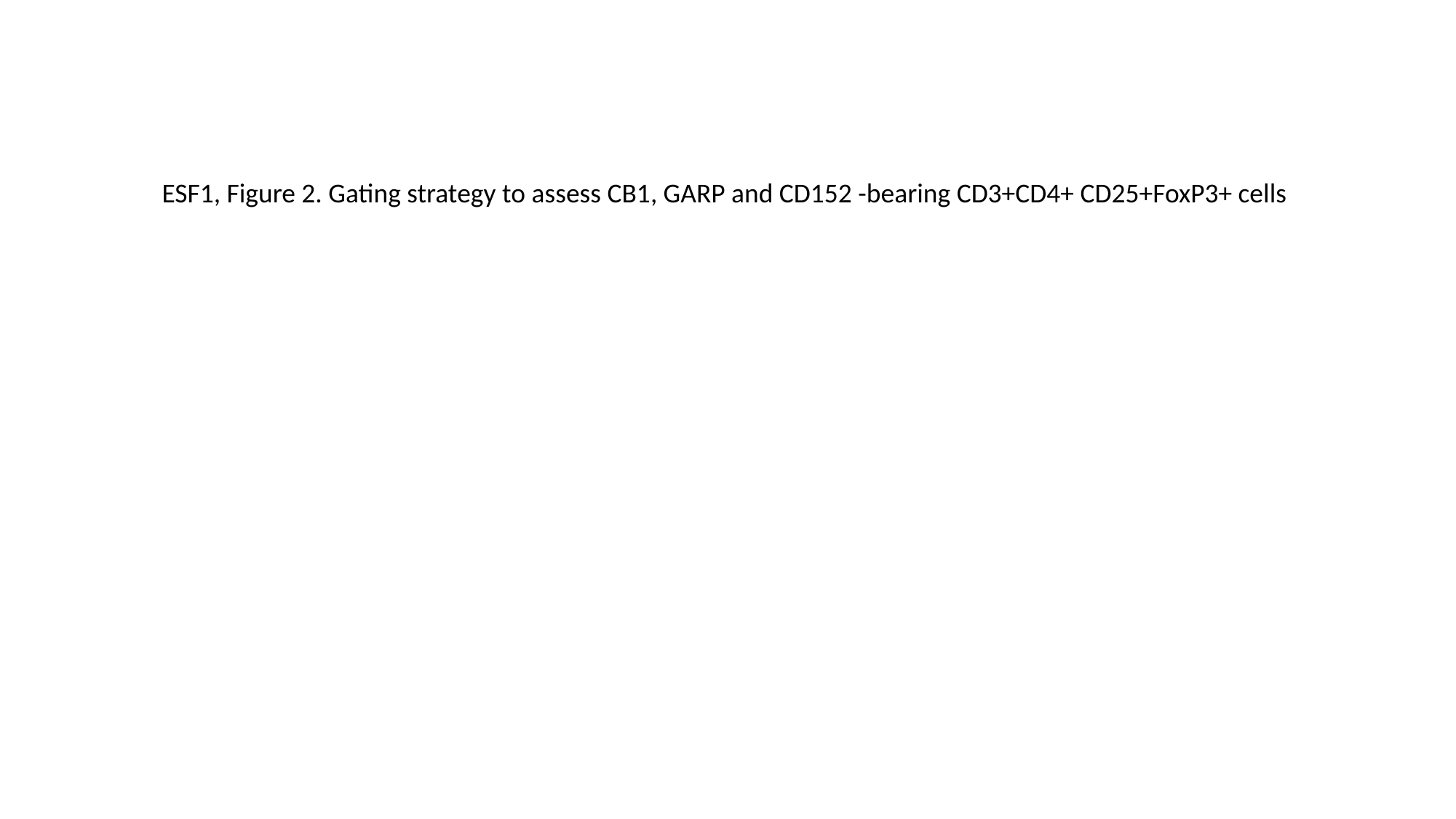

ESF1, Figure 2. Gating strategy to assess CB1, GARP and CD152 -bearing CD3+CD4+ CD25+FoxP3+ cells
