## Supplementary material for "T cell activation via the CD40 ligand and transferrin receptor and deficits in T regulatory cells are associated with major depressive disorder and severity of depression": ESF2

**ELECTRONIC SUPPLEMENTARY FILE 2 (ESF2)**

**ESF2, Table 1. T**he different CD and cell surface markers, as well as their key functions, measured in the present study.

| **CD Marker** | **Alternative names and functions** |
| --- | --- |
| CD3+ | T3, pan T-cell marker |
| CD4+ | T4, T-cell marker, T helper cell |
| CD8+ | Leu2, T-cell marker, T cytotoxic cell |
| CD69+ | Very early activation T-cell marker of especially tissue-resident immune cells (marker of tissue retention), involved in lymphocyte proliferation |
| CD71+ | Transferrin receptor, early activation T cell marker, promotes T cell proliferation |
| CD40L or CD154 | CD40 ligand, expressed on activated T cells, member of the TNF family, binds to CD40 (on CD8+ T cells, macrophages, antigen presenting cells, and B cells |
| CD3 + CD4 + CD69+ | CD69+ co-expressing CD3 CD4 cells |
| CD3 + CD8 + CD69+ | CD69+ co-expressing CD3 CD8 cells |
| CD3 + CD4 + CD71+ | Proliferating CD3 + CD4+ cells |
| CD3 + CD8 + CD71+ | Proliferating CD3 + CD8+ cells |
| CD3 + CD4 + CD40L+ | Activated CD3 + CD4+ T cells |
| CD3 + CD8 + CD40L+ | Activated CD3 + CD4+ T cells |
| CD25+FoxP3+CB1+ (gated from CD3+CD4) | CB1 bearing T regulatory (Treg) cells |
| CD25+FoxP3+CD152+ (gated from CD3+CD4) | CTLA-4 or CD152 is expressed on Treg cells, immune checkpoint that negatively regulates the immune response, CD152 bearing Treg cells |
| CD25+FOXP3+GARP+ (gated from CD3+CD4+) | Glycoprotein A repetitions predominant, TGF-β1 receptor, Treg activation marker, these Tregs release TGF-β1, promote immune tolerance |

**ESF2, Table 2.** Cytokine, chemokines and growth factors examined in the current study

| **Protein abbreviations** | **Protein name** | **Gene Symbol** |
| --- | --- | --- |
| IL-1β | Interleukin-1β | IL1B |
| IL-1RA | Interleukin-1 receptor antagonist | IL1RN |
| IL-2 | Interleukin-2 | IL2 |
| IL-4 | Interleukin-4 | IL4 |
| IL-5 | Interleukin-5 | IL5 |
| IL-6 | Interleukin-6 | IL6 |
| IL-7 | Interleukin-7 | IL7 |
| CXCL8 / IL-8 | C-X-C motif chemokine ligand 8 | CXCL8 |
| IL-9 | Interleukin-9 | IL9 |
| IL-10 | Interleukin-10 | IL10 |
| IL-12 | Interleukin-12 | IL12 |
| IL-13 | Interleukin-13 | IL13 |
| IL-15 | Interleukin-15 | IL15 |
| IL-17 | Interleukin-17 | IL17 |
| CCL11 | Eotaxin | CCL11 |
| FGF2 | Fibroblast growth factor 2, Basic fibroblast growth factor | FGF2 |
| G-CSF | Granulocyte Colony Stimulating Factor, Colony Stimulating Factor 3 (Granulocyte) | CSF3 |
| GM-CSF | Granulocyte-macrophage colony-stimulating factor, Colony-stimulating factor 2 | CSF2 |
| IFN-γ | Interferon-γ | IFNG |
| CXCL10 /IP-10 | C-X-C motif chemokine ligand 8, Interferon gamma-induced protein 10 | CXCL10 |
| CCL2 / MCP1 | C-C Motif Chemokine Ligand 2 | CCL2 |
| CCL3 / MIP-1α | Macrophage inflammatory protein-1 alpha, C-C Motif Chemokine Ligand 3 | CCL3 |
| PDGF | Platelet Derived Growth Factor Subunit B | PDGFB |
| CCL4 / MIP-1β | C-C Motif Chemokine Ligand 4, Macrophage Inflammatory Protein 1-Beta, Lymphocyte Activation Gene 1 Protein | CCL4 |
| CCL5 /RANTES | C-C Motif Chemokine Ligand 5, Regulated Upon Activation, Normally T-Expressed, And Presumably Secreted | CCL5 |
| TNF-α | Tumor Necrosis Factor-Alpha | TNF |
| VEGF | Vascular Endothelial Growth Factor | VEGFA |

**ESF2, Table 3**. Description of the immune profiles used in this study

| **Immune Profile** | **Members** |
| --- | --- |
| **M1 macrophage** | IL-1β, sIL-1RA, IL-6, TNF-α, CXCL8, CCL3 |
| **T helper-1** | IL-2, IFN-γ, IL-12 |
| **IRS** | IL-1β, IL-6, TNF-α, CXCL8, CCL3, IL-2, IFN-γ, IL-12, IL-17, IL-15, G-CSF, GM-CSF, CXCL10, CCL5, CCL2 |
| **Neurotoxicity** | IL-1β, IL-6, TNF-α, CXCL8, CCL3, IL-2, IFN-γ, IL-12, IL-17, CXCL10, CCL11, CCL5, CCL2 |
| **T cell growth** | IL4, IL9, IL12, GM-CSF, IL-15 |

IRS: immune-inflammatory response system

**ESF2, Table 4.** Measurements of the frequencies of T cell activation markers

in baseline (UNSTIM) and stimulated (STIM) conditions in major depressive

disorder (MDD) and controls (HC).

| **Estimates** | | | | | |
| --- | --- | --- | --- | --- | --- |
| MDD | UN_STIM | Mean | Std. Error | 95% Wald Confidence Interval | |
|  |  |  |  | Lower | Upper |
| HC | UNSTIM | 4.8226 | 1.59616 | 1.6942 | 7.9510 |
|  | STIM | 50.4121 | .96969 | 48.5115 | 52.3126 |
| MDD | UNSTIM | 6.2336 | 1.54829 | 3.1990 | 9.2682 |
|  | STIM | 52.5532 | 1.21500 | 50.1719 | 54.9346 |

Results are shown as estimated marginal mean (in percentage) and SE.

**ESF2, Table 5.** Differences in unstimulated (UNST) and stimulated (STIM) changes of median immune fluorescence (MFI) of T lymphocytes in healthy controls (HC) and major depressed patients (MDD)

|  | **Condition** | **Diagnosis** | | **Tests of Model effects** | | |
| --- | --- | --- | --- | --- | --- | --- |
| **MFI (z scores)** |  | **HC ^a^**  **n=20** | **MDD ^c^**  **n=30** | **Effects** | **Wald**  **df=1** | **p** |
| CD3+CD69+ | UNST | -0.665 (0.105) | -0.785 (0.2) | Group (G)  Gxtime (T) | 0.047  0.779 | 0.829  0.371 |
|  | STIM | 0.702 (0.06) | 0.761 (0.063) |  |  |  |
| **CD3+CD71+** | UNST | -1.109(0.12) ^c^ | -0.791(0.088) ^a^ | G | 4.222  4.024 | **0.040**  **0.045** |
|  | STIM | 0.895(0.037) | 0.931(0.035) | GXT |  |  |
| CD3+CD154+ | UNST | -1.001 (0.098) | -0.889 (0.068) | G | 0.336  0.98 | 0.562  0.332 |
|  | STIM | 0.952 (0.049) | 0.921 (0.06) | GXT |  |  |
| CD3+HLADR+ | UNST | -0.992 (0.131) | -0.761 (0.131) | G | 3.774  0.613 | 0.052  0.434 |
|  | STIM | 0.807 (0.07) | 0.882 (0.049) | GXT |  |  |
| CD4+CD69+ | UNST | -0.843 (0.110) | -0.886 (0.127) | G | 0.266  1.469 | 0.606  0.225 |
|  | STIM | 0.776 (0.064) | 0.929 (0.06) | GXT |  |  |
| **CD4+CD71+** | UNST | -1.109 (0.116) ^c^ | -0.79 (0.085) ^a^ | G | 3.88  6.144 | **0.049**  0.013 |
|  | STIM | 0.928 (0.039) | 0.908 (0.045) | GXT |  |  |
| CD4+CD154+ | UNST | -1.013 (0.095) | -0.889 (0.063) | G | 0.153  1.882 | 0.695  0.170 |
|  | STIM | 0.981 (0.05) | 0.909 (0.058) | GXT |  |  |
| **CD4+HLADR+** | UNST | -1.13 (0.138) ^c^ | -0.633 (0.119) ^a^ | G | 8.42  3.973 | **0.004**  0**.046** |
|  | STIM | 0.754 (0.078) | 0.879 (0.077) | GXT |  |  |
| CD8+CD69+ | UNST | -0.71 (0.293) | -0.56 (0.143) | G | 0.027  0.861 | 0.870  0.353 |
|  | STIM | 0.667 (0.086) | 0.587 (0.092) | GXT |  |  |
| CD8+CD71+ | UNST | -0.955 (0.093) | -0.928 (0.064) | G | 0.807  0.648 | 0.369  0.421 |
|  | STIM | 0.867 (0.065) | 0.986 (0.05) | GXT |  |  |
| CD8+CD154+ | UNST | -0.907 (0.099) | -0.94 (0.068) | G | 1.017  1.874 | 0.313  0.171 |
|  | STIM | 0.82 (0.057) | 0.997 (0.066) | GXT |  |  |
| CD8+HLADR+ | UNST | -0.919(0.128) | -0.855(0.116) | G | 0.148  0.081 | 0.700  0.766 |
|  | STIM | 0.873(0.062) | 0.886(0.048) | GXT |  |  |
| CD25+ FOXP3+CB1+ | UNST | -0.025 (0.160) | -0.376 (0.108) | G | 1.047  0.452 | 0.306  0.501 |
|  | STIM | 0.343 (0.29) | 0.168 (0.214) | GXT |  |  |
| CD25+ FOXP3+CD152+ | UNST | -0.43 (0.235) | -0.817 (0.171) | G | 1.865  1.229 | 0.172  0.268 |
|  | STIM | 0.728 (0.095) | 0.622 (0.059) | GXT |  |  |
| CD25+ FOXP3+GARP+ % | UNST | -0.717(0.279) | -0.742(0.112) | G | 0.052  0.218 | 0.820  0.640 |
|  | STIM | 0.669(0.074) | 0.773(0.057) | GXT |  |  |

**ESF2, Table 6**. Measurements of the median fluorescence intensity of T regulatory cells

in baseline (UNST) and stimulated (STIM) conditions in major depressive

disorder (MDD) and healthy controls (HC).

| **Estimates** | | | | | |
| --- | --- | --- | --- | --- | --- |
| Group | UNST_STIM | Mean | Std. Error | 95% Wald Confidence Interval | |
|  |  |  |  | Lower | Upper |
| HC | UNST | -.1969023 | .09951962 | -.3919572 | -.0018475 |
|  | STIM | .3848233 | .16977562 | .0520692 | .7175774 |
| MDD | UNST | -.4500016 | .05696057 | -.5616423 | -.3383610 |
|  | STIM | .3268810 | .10627718 | .1185816 | .5351805 |

Results are shown as estimated marginal mean (in percentage) and SE.
